# Evaluating the impact of a policy restricting reimbursement of lidocaine plaster prescribing on opioid and other analgesic prescribing

**DOI:** 10.64898/2026.09.10.26362704

**Authors:** Molly Mattsson, Ahmed Hassan Ali, Brian MacKenna, Michelle Flood, Ciara Kirke, Emma Wallace, Tom Fahey, Fiona Boland, Frank Moriarty

## Abstract

**Background:** Introducing a reimbursement restriction on lidocaine plaster prescribing in Ireland may have had unintended effects on prescribing of other analgesics with misuse potential. This study evaluates the impact of this policy on use, treatment intensification and initiation of opioids, gabapentinoids, and other analgesics.

**Methods:** This cohort study included adults eligible for Ireland’s public health cover (approximately 33% of the population), using primary care dispensing data. The policy was fully implemented in December 2017, requiring individual patient approval for coverage of lidocaine plasters. In interrupted time series analysis, we examined change in level and trend of prescribing of other analgesics. In cohort analysis we compared lidocaine users to users of any other analgesic pre-policy and assessed outcomes of treatment intensification and initiation of opioids, gabapentinoids, and other analgesics in the three months post-policy.

**Results:** Among 1.65 million individuals, up to 1.4% were dispensed lidocaine plasters monthly pre-policy. Population-level use of other analgesics remained largely unchanged (except for topical agents). Compared to other analgesic users, lidocaine plasters users were significantly more likely to experience treatment intensification (adjusted risk ratio 1.47, 95%CI 1.38-1.57) of opioids after the policy, and initiation of strong (2.07, 1.85-2.30) and weak opioids (1.38, 1.21-1.59), controlling for baseline differences. Similar results were identified for gabapentinoid treatment intensification (1.26, 1.16-1.38) and initiation (1.34, 1.11-1.61), as well as for other analgesics.

**Conclusions:** This policy aimed at reducing low-value care was associated with greater opioid and gabapentinoid consumption, exposing patients to greater risk of drug-related harm from analgesic use and misuse.

## Background

Potentially inappropriate prescribing, where the actual or potential harms of therapy outweigh the benefits^1^, is a substantial challenge to healthcare systems globally. One important aspect of inappropriate prescribing is low-value care, typically defined as health services that provide little or no benefit, has potential to cause harm, incur unnecessary costs, or waste limited healthcare resources.^2,3^ Strategies to promote appropriate prescribing and ensure safe, effective, and cost-effective medicines use may be grouped broadly as targeted or system-oriented approaches, with targeted approaches comprising educational and managerial interventions aimed at prescribers and users and system-oriented strategies, including regulatory and economic interventions aiming to restrict prescribing^4^. Both approaches may have unintended consequences, such as an increase in use of alternative medications or inadequate access to medications. A systematic review on the effect of formulary restrictions on patient and payer outcomes found that formulary coverage decisions may provide benefits like lower drug utilisation and pharmacy cost savings; but with unintended consequences like reduced medication adherence and treatment satisfaction and increased healthcare utilisation and medical costs^5^. Careful evaluation of the consequences of prescribing restrictions, both before policy implementation and in the form of continued re-evaluation after implementation, is therefore warranted.

In Ireland, the Health Service Executive (HSE) introduced a restricted reimbursement system for lidocaine 5% plasters in 2017, requiring prior authorisation. Lidocaine 5% plasters (marketed as Versatis® in Ireland) are licensed for the indication of symptomatic relief of neuropathic pain associated with previous herpes zoster infection, known as post-herpetic neuralgia (PHN), in adults, with treatment to be re-evaluated after 2-4 weeks and discontinued if the desired outcomes are not achieved^6^. Although the licensed therapeutic indication of lidocaine 5% plasters is limited to PHN, the HSE expenditure on this medicine (over €30 million with over 25,000 or 1.5% of individuals dispensed this medicine on the main public health scheme in 2016^7^) suggested that there was considerable off-label use. Evidence does not support its use in wider indications, with only non-steroidal anti-inflammatory drugs and capsaicin as the main topical analgesic drugs recommended in NICE guidelines for osteoarthritic and neuropathic pain^8,9^. An overview of Cochrane reviews found that the evidence for lidocaine plasters in the treatment of acute or chronic pain was of very low quality^10^. From 1^st^ September 2017, the HSE introduced an online reimbursement application system for lidocaine plasters, where prescribers needed to indicate the antiviral prescribed for the herpes zoster infection, thus ensuring that lidocaine plasters were not prescribed for other indications. If not submitted through the reimbursement system, lidocaine plasters would no longer be reimbursed through community drug schemes. This applied to new users from 1^st^ September 2017, and all users from 1^st^ December 2017.

Previous research has shown that the introduction of the online reimbursement system was associated with an immediate reduction in prescribing of lidocaine plasters by 97.2% and a reduction in public expenditure from €27 million in 2016 to €2 million in 2018^11,12^. However, little is known regarding how this change in lidocaine plaster prescribing may have influenced other analgesic prescribing, including switching to or intensification of opioids, gabapentinoids, and NSAIDs, all of which have been associated with serious adverse effects, including misuse and dependency for opioids and gabepentinoids, and adverse cardiovascular events and gastrointestinal bleeding for NSAIDs^13,14^.

The aim of this study was to investigate the impact of lidocaine reimbursement restrictions on analgesic use by patients in Ireland. This was addressed through the following specific objectives:

1. To evaluate how the introduction of lidocaine reimbursement restrictions impacted the level and trend of overall analgesic use in the GMS population.
2. To evaluate how initiation and dose escalation for analgesics following the introduction of lidocaine reimbursement restrictions differed between existing users of lidocaine plasters and existing users of other analgesics.

## Methods

This is a cohort study, incorporating an interrupted time series analysis. It forms part of a larger project focused on analgesic and sedative prescribing, the protocol for which has been previously published^15^. The study was approved by the RCSI University of Medicine and Health Sciences Research Ethics Committee (ref: REC202201015), the Health Service Executive (HSE) Reference Research Ethics Committee B (ref: RRECB1022FM), and the Health Research Consent Declaration Committee (22-008-AF1).

### Population and data sources

This study included individuals eligible for the General Medical Services (GMS) scheme in Ireland and was obtained from the Health Service Executive (HSE) Primary Care Reimbursement Service (PCRS), which administers community drug schemes in Ireland. Eligibility to this scheme is based on age and income and covers approximately 32% of the population, and therefore eligible persons tend to be older and more socioeconomically deprived than the general population^16^. Pharmacies transmit claims for medications dispensed under the GMS scheme to the PCRS at the end of each month for reimbursement and the PCRS pharmacy claims database therefore contains records of all prescribed medications which were dispensed to individuals eligible for the scheme.

Data were requested from the PCRS for this study through submission of an information request, in line with the PCRS privacy notice (https://www.mymedicalcard.ie/privacy-statement) and procedures, and data were provided to the research team under a data transfer agreement. Data were at the level of individual medications dispensed to each patient. For each item, information included anonymised identifiers for individuals, prescribers and pharmacies, Local Health Office (LHO) where the drug was dispensed, date of dispensing, sex and age group of the individual, drug information (World Health Organisation (WHO) Anatomical Therapeutic Chemical (ATC) code, product name and strength, quantity dispensed, and cost.

### Drugs

Analgesics were identified using WHO ATC codes. Outcomes were derived for drug classes, including opioids, gabapentinoids, systemic non-steroidal anti-inflammatory drugs (NSAIDs), topical NSAIDs, and lidocaine plasters, topical capsaicin and paracetamol as individual drugs.^17^ Defined daily doses (DDD) were applied to relevant ATC codes by route of administration^18^. The WHO DDD, defined as “the assumed average maintenance dose per day for a drug used for its main indication in adults”^19^ was used for oral NSAIDs. For opioids, oral morphine equivalents (OME) were used.^20^

### Outcomes

To address the study aim, multiple outcomes were derived for the drug classes outlined above, and those considered for objective 1 were prevalence of dispensing for each class, as well as prevalence of initiations, discontinuations, and chronic use. These outcomes were calculated for each month from January 2014 to December 2019. Denominators for rates were obtained from PCRS annual reports which report number of eligible individuals at the end of each calendar year^21^. Prevalence of use was calculated as the proportion of individuals dispensed a relevant medicine within each month. Initiation was defined as a dispensing to an individual with no dispensing in the previous 90 days, and discontinuation as an individual with no dispensing in the 90 days following a dispensing. Long-term use was derived for >90 days.^22^

For objective 2, the outcomes considered were treatment intensification, initiation, and total standard doses dispensed during the three months post-policy implementation for opioids, gabapentinoids, systemic NSAIDs and paracetamol. Treatment intensification was defined as a month-on-month increase in OMEs (opioids) or DDDs (other analgesics) dispensed to an individual in any of the three months following the introduction of the reimbursement restriction (December 2017–February 2018). Therefore, this captures dose increases and initiations.

### Analysis

To evaluate how the introduction of lidocaine reimbursement restrictions impacted the level and trend of overall prevalence of dispensing, prevalence of initiations, discontinuations, and chronic use of analgesic drug classes, segmented regression models were fitted. The effects of the two stages of the restrictions (new users in September 2017 and existing users in December 2017) were both included.

Each segmented regression model was parameterised to estimated four elements:

i. the rate at the beginning of the study period (i.e., the model intercept),
ii. the trend prior to the intervention,
iii. immediate change in rate from pre to post intervention, and
iv. change in trend over time from pre to post intervention.

The analysis for objective 2 evaluated how the prevalence of initiations and treatment intensification of opioids, systemic NSAIDs, paracetamol, and gabapentinoids in the three months following the introduction of reimbursement restrictions (December 2017-February 2018) differed between regular users of lidocaine plasters November 2017 (at least three dispensings in the preceding 90 days) and regular users of other analgesics. Generalised linear regression models (with binomial distribution and log link), adjusting for other analgesic use, current dose of the drug of interest, age, and sex, were used, to estimate risk ratios (RR) with 95% confidence intervals (CIs). Initiations of strong and weak opioids were examined separately. The number needed to treat (NNT) was calculated for treatment intensification and initiation, i.e. the number of lidocaine users who would need to be exposed to the policy for one additional patient to have treatment intensification or initiation of another analgesic. This was calculated by estimating the absolute risk increase by the formula *Event rate* (*comparator group*) x (1 − *adjusted risk ratio*), and taking the reciprocal of this. NNTs should be interpreted as descriptive measures of absolute risk difference rather than causal treatment effects.

To assess how total OMEs for opioids and DDDs for gabapentinoids, systemic NSAIDs, and paracetamol in the three months post-restriction differed between existing lidocaine users and other analgesic users, negative binominal regression models, adjusting for other analgesic use, current dose of the drug of interest, age, and sex, were used, to estimate incidence rate ratios (IRR) and 95% CIs.

Multivariate regression analyses were also stratified by age group (i.e. aged <65 years, and ≥65 years) to assess the robustness of findings across both adults and older adults. Further sensitivity analysis included a time period control, including those eligible as lidocaine or other analgesic users in November 2016 and assessing outcomes in the period December 2016-February 2017. Regression analyses were repeated with the addition of the 2016 cohort, adjusting standard error for clustering of any repeated observations of the same individuals, with the interaction between lidocaine group and time period interpreted as the main parameter of interest. A final sensitivity analysis repeated the primary analysis comparing individuals with any lidocaine plaster dispensing in November 2017 to those with any other analgesic dispensing (rather than chronic use). All analysis was conducted using Stata version 18 (College Station, TX: StataCorp LLC) statistical significance was assumed at p<0.05. For the interrupted time-series analysis, the ITSA package was used^23^.

## Results

Prevalence of lidocaine plaster dispensing dropped by 0.46 percentage points (pp) in September 2017, a monthly decline of 0.18 pp from September to November 2017, and a further 0.42 pp drop in December 2017 (see Figure 1, Supplementary Table 1). A similar pattern was observed for the prevalence of chronic use, both driven by a reduction in lidocaine initiations in September 2017 (0.32 pp relative to August 2017) and to a lesser extent in December 2017 (0.03 pp relative to November 2017), and an increase in discontinuations in December 2017 (reaching 0.28%) and January 2018 (0.17%).

**Figure 1.**
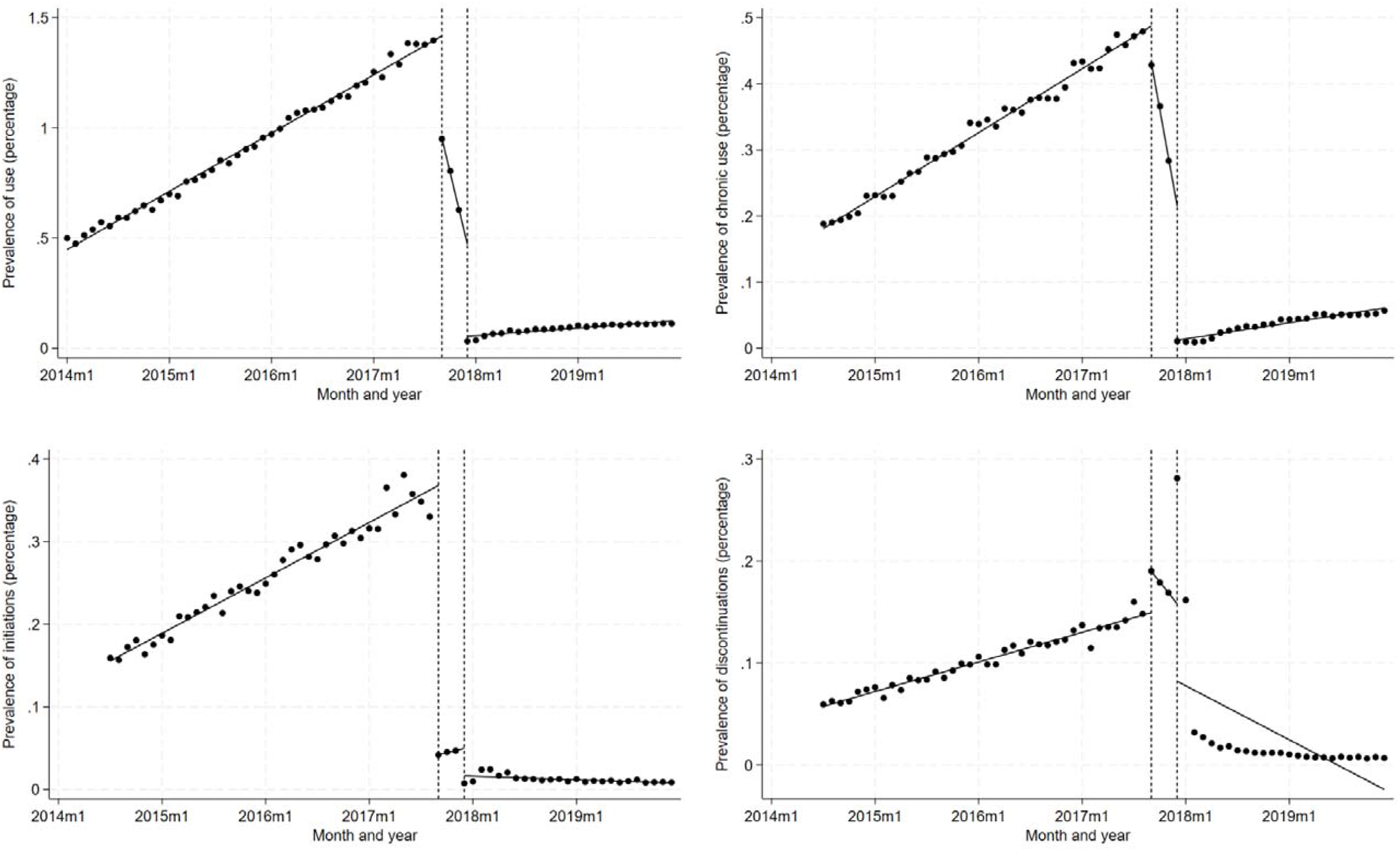
Interrupted time series analysis for prevalence, chronic use, initiations, and discontinuations of lidocaine plasters.

Considering alternative agents, overall prevalence of dispensing of topical NSAIDs in the GMS scheme population increased significantly in September 2017 (0.11 pp) as did topical capsaicin (0.02 pp), which also saw increases in the monthly trend, however this decreased significantly after December 2017. For topical capsaicin, this was mainly driven by initiations in September 2017 (0.01 pp increase, see Supplementary Figure 1, Supplementary Table 2), and an increasing trend of chronic use between September 2017 and November 2017 (see Supplementary Figure 2, Supplementary Table 2).

There were no overall significant changes in prevalence of systemic NSAIDs, opioids, gabapentinoids or paracetamol in the overall GMS population (Figure 2, Supplementary Table 2). Notably, there was an overall drop in systemic NSAID and opioid initiations in September 2017 (0.014 pp and 0.07 pp respectively). For chronic use of oral drugs, there was little change over time however all had small, significant drops in September 2017 (e.g. 0.04 pp for systemic NSAIDs) offset by similar, generally significant increases in December 2017 (e.g. 0.06 pp for systemic NSAIDs) (see Supplementary Figure 2, Supplementary Table 2).

**Figure 2.**
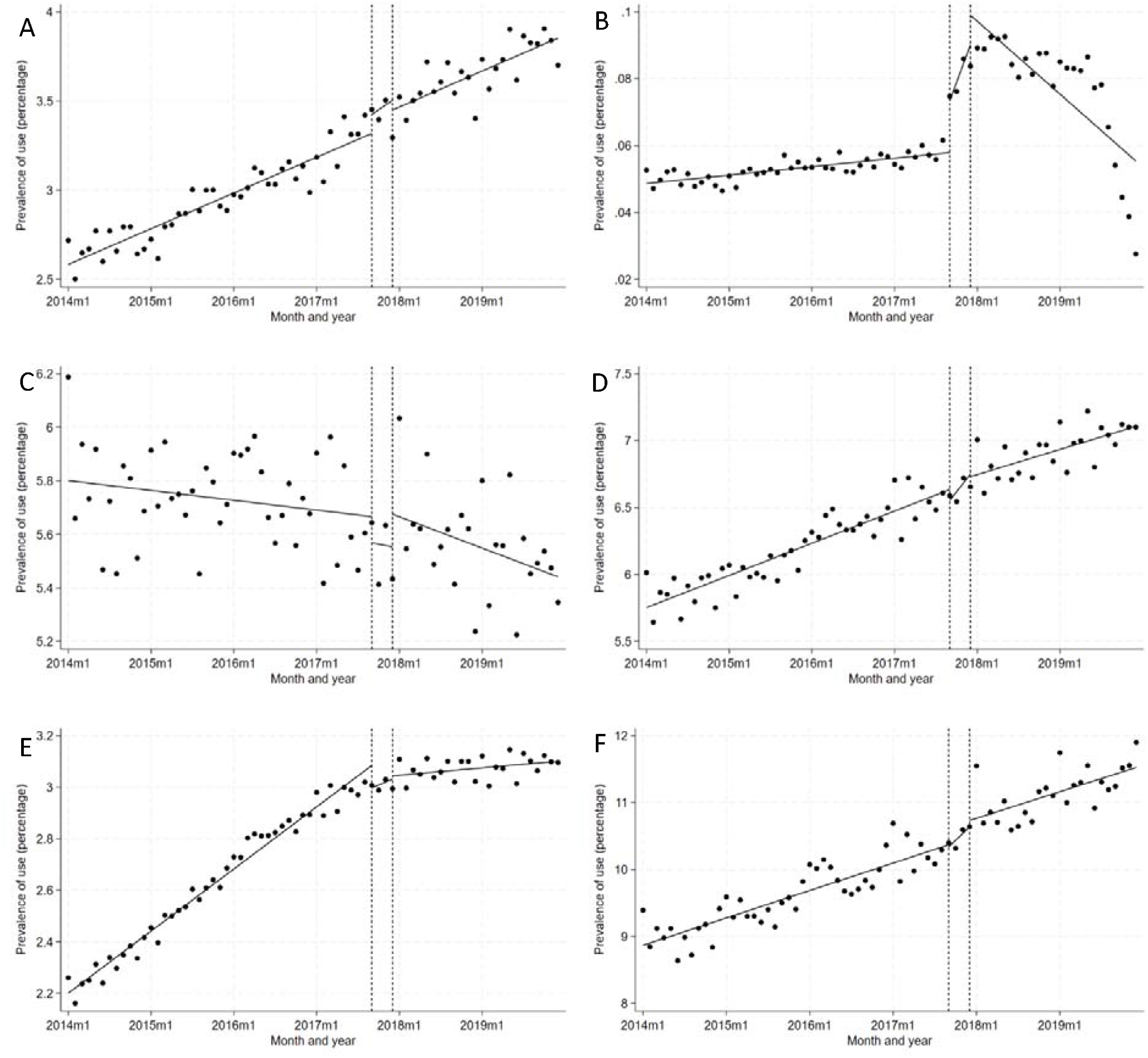
Interrupted time series analysis for prevalence of use of (a) topical NSAIDs, (b) topical capsaicin, (c) systemic NSAIDs, (d) opioids, (e) gabapentinoids, and (f) paracetamol.

Comparing regular lidocaine users to regular users of other analgesic users in November 2017 (without regular lidocaine plaster use) before implementation of restriction for existing users, lidocaine users were older, more often female, had higher prevalence of use of other analgesic drug classes (except for systemic NSAIDs) as well as higher prevalence of chronic analgesic use. They also tended to be prescribed a higher number of standard doses for drug classes, except for paracetamol and gabapentinoids.

Considering the primary outcomes, treatment intensification (a higher standard dose dispensed in any of the three months following the restriction introduction versus November 2017) was more common among lidocaine users (prevalence of 14.6-39.9%) compared to other analgesic users for all drug classes except systemic NSAIDs. Similar was true for initiations, with higher prevalence among lidocaine users (from 4.0 to 34.9% across systemic analgesic drug classes), except for weak opioid and systemic NSAID initiations. Among users of each drug class, there were higher numbers of standard doses dispensed monthly in the lidocaine group versus the other analgesic group.

In generalised linear model regression analysis, after adjusting for age group, sex, and other analgesics medication use, lidocaine users had significantly higher risk of intensification for opioids (adjusted RR 1.47, 95%CI 1.38-1.57), gabapentinoids (1.26, 95%CI 1.16-1.38) as well as systemic NSAIDs and paracetamol. This equates to a NNT of 7.5 (95%CI 6.2.-9.3) for one individual to experience opioid intensification, and 32 (95%CI 22-52) for gabapentinoids (see Supplementary Table 3). Results were similar in adjusted analysis of initiations, with higher risk of initiation for strong opioids (adjusted risk ratio 12.07, 95%CI 1.85-2.30), weak opioids (1.38, 95%CI 1.21-1.59), and gabapentinoids (1.34, 95%CI 1.11-1.61). Risk ratios were of similar magnitude for paracetamol and were smaller for systemic NSAIDs. The NNT for strong opioid initiation was 11.5 (95%CI 9.5-14.5), for weak opioids was 38 (95%CI 53-69) and 84 (95%CI 47-260) for gabapentinoids (see Supplementary Table 3). Considering results stratified by age group, associations were of stronger magnitude for gabapentinoid intensification and initiation among older adults compared to those aged <64 years, while for systemic NSAID treatment intensification and initiation, these association were weaker among older adults (see Supplementary Table 4). In sensitivity analysis further adjusting for treatment period effects, comparing to the difference between these groups of patients one year earlier, the associations between lidocaine plaster use and outcomes were of a smaller magnitude for opioids, gabapentinoids and paracetamol (with null, non-significant results for gabapentinoid treatment intensification and initiations), while the magnitude of association for systemic NSAIDs increased (Supplementary Table 5). In a further sensitivity analysis comparing individuals with any lidocaine plasters dispensing in November 2017 to those with any dispensing of another analgesic, compared to the primary analysis, associations remained statistically significant and of similar magnitudes (Supplementary Table 6).

Among those with opioid use post-restriction implementation, those in the lidocaine group had a significantly higher rate of standard opioid doses (i.e. OMEs) compared to the other analgesic group after adjustment (adjusted IRR 1.13, 95%CI 1.09-1.17). Similarly, the lidocaine group had significantly higher rates of systemic NSAID and paracetamol standard doses, however there was no significant difference in the rate of standard doses for gabapentinoids. Comparing age groups, the association with higher rates of standard doses for opioids were stronger among adults aged 64 or less compared to older adults (see Supplementary Table 4). In sensitivity analysis adjusting for November 2016 prescribing, opioid and paracetamol associations were non-significant (Supplementary Table 5).

## Discussion

This study found that at the population level, only topical NSAIDs and capsaicin had statistically significant increases in use following the introduction of a reimbursement restriction involving prior authorisation for lidocaine 5% plasters in Ireland (though with a very small absolute increase for a period of approximately 18 months), while there was no major change in level or trend of systemic analgesic prescribing. This likely reflects the relatively low prevalence of lidocaine plaster use, lidocaine plasters were dispensed to fewer than 1.5% of the GMS population, and high proportions of those dispensed lidocaine plasters were already prescribed systemic analgesics (see Table 1). However, at the individual patient level, those dispensed lidocaine plasters had significantly higher risk of having treatment intensification or being initiated on each of the systemic analgesic drug classes we examined after the restriction on reimbursement. In particular for opioids, there was a 47% increased risk of treatment intensification, and 38% and 107% increased risk of initiating a weak or strong opioid respectively. This suggests those individuals who were no longer able to access lidocaine plasters at low cost through public health cover required further analgesic treatment. Compared to a previous evaluation of the reimbursement restriction policy in Ireland using the same data source, our study identified apparent switching to a range of analgesic treatments, where the previous study only evaluated use of recommended alternatives (topical NSAIDs and capsaicin), and identified 30% of pre-implementation lidocaine plaster users who initiated these topical agents in the following 6 months.^12^ In contrast, we found prevalence of initiation of 4.0-34.9% among systemic drug classes, and prevalence of treatment intensification of 14.6-39.9%, indicating a substantial impact of this policy change not previously examined.

**Table 1.** Characteristics of GMS eligible individuals dispensed analgesics in November 2017.

|  | Lidocaine users (n=4,563) | Other analgesic users (n=168,015) |
| --- | --- | --- |
| **Age, mean ± SD** | 73.2 ± 14.5 | 65.9 ± 16.8 |
| **Female sex, % (n)** | 74.8 (3,414) | 64.5 (108,318) |
| **Prevalence of any medication use, % (n)** |  |  |
| Opioids | 53.6 (2,447) | 45.9 (77,194) |
| Gabapentinoids | 32.5 (1,483) | 24.6 (41,256) |
| Systemic NSAIDs | 17.7 (809) | 26.7 (44,786) |
| Paracetamol | 67.8 (3,093) | 63.8 (107,258) |
| Topical analgesic | 30.5 (1,393) | 22.7 (38,166) |
| Topical NSAIDs | 29.8 (1,361) | 22.3 (37,400) |
| Topical capsaicin | 1.1 (51) | 0.6 (1,066) |
| **Prevalence of chronic use (90 days), % (n)** |  |  |
| Opioids | 45.1 (2,059) | 29.1 (48,817) |
| Gabapentinoids | 29.3 (1,336) | 16.4 (27,625) |
| Systemic NSAIDs | 11.8 (538) | 10.3 (17,319) |
| Paracetamol | 55.8 (2,546) | 38.9 (65,399) |
| Topical analgesic | 20.4 (931) | 8.6 (14,456) |
| Topical NSAIDs | 20.1 (915) | 8.5 (14,279) |
| Topical capsaicin | 0.5 (22) | 0.1 (217) |
| **Standard daily doses (median, IQR)** |  |  |
| Opioids | 840 (308, 1,740) | 450 (144, 1,120) |
| Gabapentinoids | 16 (9.3, 30) | 14 (8.8, 28) |
| Systemic NSAIDs | 30 (20, 56) | 28 (15, 45) |
| Paracetamol | 20 (16.7, 33.3) | 16.7 (10, 20) |
| **Individuals with above median standard doses, % (n)** |  |  |
| Opioids | 35.6 (1,625) | 22.2 (37,293) |
| Gabapentinoids | 17.2 (786) | 11.8 (19,844) |
| Systemic NSAIDs | 10.2 (463) | 13.1 (21,949) |
| Paracetamol | 40.8 (1,863) | 27.1 (45,496) |

IQR = interquartile range; NSAID = non-steroidal anti-inflammatory drug; SD = standard deviation

**Table 2.** Treatment intensifications, initiations, and average monthly standard doses between December 2017 and February 2018 in GMS eligible individuals dispensed analgesia in November 2017.

|  | Lidocaine users | Other analgesic users |
| --- | --- | --- |
| **Treatment intensification, % (n)** |  |  |
| Opioids | 35.9 (1,638) | 28.3 (47,500) |
| Gabapentinoids | 14.6 (664) | 12.1 (20,243) |
| Systemic NSAIDs | 17.4 (793) | 20.1 (33,771) |
| Paracetamol | 39.9 (1,819) | 34.7 (58,227) |
| **Initiations, % (n)\*** |  |  |
| Strong opioids | 14.4 (404) | 8.1 (10,037) |
| Weak opioids | 6.7 (238) | 6.9 (8,873) |
| Gabapentinoids | 4.0 (123) | 3.5 (4,415) |
| Systemic NSAIDs | 11.9 (447) | 13.6 (16,785) |
| Paracetamol | 34.9 (513) | 25.0 (15,166) |
| **Monthly standard doses (median, IQR)** |  |  |
| Opioids | 800 (300, 1,680) | 426 (107, 1,066) |
| Gabapentinoids | 15 (8, 28) | 14 (7, 28) |
| Systemic NSAIDs | 30 (15, 49) | 28 (12, 42) |
| Paracetamol | 22 (17, 33) | 17 (12, 25) |

\* Expressed as a percentage of non-users of the outcome drug in November 2017 (i.e. those who could initiate the outcome drug in the period December 2017-February 2018). IQR = interquartile range; NSAID = non-steroidal anti-inflammatory drug

**Table 3.** Generalised linear model regression results for individuals dispensed lidocaine in November 2017 versus individuals dispensed other analgesics.

|  | Unadjusted RR (95% CI) | Adjusted RR (95% CI) |
| --- | --- | --- |
| **Treatment intensification** |  |  |
| Opioids | 1.42 (1.34-1.51) | 1.47 (1.38-1.57) |
| Gabapentinoids | 1.24 (1.14-1.35) | 1.26 (1.16-1.38) |
| Systemic NSAIDs | 0.84 (0.77-0.90) | 1.12 (1.04-1.21) |
| Paracetamol | 1.25 (1.18-1.33) | 1.35 (1.27-1.44) |
| **Initiations** |  |  |
| Strong opioids | 1.38 (1.25-1.53) | 2.07 (1.85-2.30) |
| Weak opioids | 0.84 (0.74-0.96) | 1.38 (1.21-1.59) |
| Gabapentinoids | 1.00 (0.85-1.18) | 1.34 (1.11-1.61) |
| Systemic NSAIDs | 0.76 (0.69-0.84) | 1.13 (1.02-1.26) |
| Paracetamol | 0.97 (0.89-1.06) | 1.66 (1.49-1.86) |

CI = confidence interval; NSAID = non-steroidal anti-inflammatory drug, RR = risk ratio. Models adjusted for age group, sex, and covariates for each non-outcome medication use, chronic use, and above median standard dose use in November 2017. Treatment intensification models also adjusted for November 2017 standard doses of outcome medication.

**Table 4.** Negative binomial regression results for monthly standard doses dispensed between December 2017 and February 2018 for individuals dispensed lidocaine in November 2017 versus individuals dispensed other analgesics (restricted to individuals dispensed the drug of interest)

|  | Unadjusted IRR (95% CI) | Adjusted IRR (95% CI) |
| --- | --- | --- |
| **Standard daily doses** |  |  |
| Opioids | 1.33 (1.27-1.40) | 1.13 (1.09-1.17) |
| Gabapentinoids | 1.05 (1.00-1.09) | 0.99 (0.96-1.02) |
| Systemic NSAIDs | 1.00 (0.95-1.04) | 1.03 (1.00-1.07) |
| Paracetamol | 1.16 (1.13-1.18) | 1.07 (1.06-1.09) |

CI = confidence interval, IRR = incidence rate ratio, NSAIDs = non-steroidal anti-inflammatory drugs. Models adjusted for age group, sex, and covariates for each non-outcome medication use, chronic use, and above median standard dose use, and standard doses of outcome medication in November 2017.

Aside from the previous evaluations of Ireland’s policy and the introduction of guidance recommending against routine use in England, reimbursement restrictions or similar strategies for lidocaine plasters do not appear to have been reported in the literature. However evaluations of policies to reduce use of other analgesics have been conducted, focusing on legal and other restrictions to opioids, gabapenintoids, and NSAIDs in the United States of America (USA). However, with the exception of opioids, unintended consequences do not appear to have been evaluated. Opioid restriction policies may have unintended consequences in increasing illicit drug use and associated harms,^24^ including heroin deaths.^25^ This is supported by qualitative evidence, where prescription opioid restrictions led to faster progress from prescription opioids to illicit heroin or fentanyl.^26^ This is in addition to the unintended consequence of reduced access to clinically effective therapy for those who may benefit from it.^27^ Further evidence on unintended consequences impacting on prescribed medications showed a law limiting opioid prescribing in Florida led to a 6% increase in gabapentinoid users and 11% in gabapentinoid prescriptions, though no significant increases in the use of muscle relaxants, benzodiazepines,^28^ or oral NSAIDs.^29^ Although the context of this research from the USA during the period of the opioid epidemic is not directly comparable to Ireland, it does provide evidence on the potential for important unintended consequences of restrictions on prescribed analgesics.

We identified higher risk of treatment intensification and initiation among individuals dispensed lidocaine plasters post-restriction implementation compared to other analgesic users. Although the substantial decrease in lidocaine plaster utilisation after restriction to licensed indications indicates extensive off-label use, the initiation and intensification of other analgesics indicates a genuine need for additional pain treatment for patients. So while the patterns observed may indicate appropriate treatment of need, most of the systemic analgesics examined carry substantial risk of medication-related harm. This includes risk of falls and fractures, sedation, and dependence with opioids and gabapentinoids,^30,31^ while systemic NSAIDs can increase risk of gastrointestinal bleeds, cardiovascular events, and acute kidney injury.^32,33^ Of note, the increase in NSAID use was primarily driven by prescribing in those aged under 65 years. This suggests prescribers may have avoided these medicines in older adults where they may be a higher risk of medication-related harm.^22^

Availability of therapeutic alternatives is an important consideration for both system-orientated or more targeted interventions to enhance prescribing. Unintended consequences can manifest as prescribing of off-label and/or potentially harmful drugs (as in the current study) or lack of intervention effectiveness (e.g. where efforts to reduce high-risk benzodiazepine use may be limited by lack of non-pharmacological options).^34^ This lack of available non-pharmacological alternatives is a particularly important in the population included in this study. The older, more socioeconomically disadvantaged cohort included in this study are more likely to live with multiple long-term conditions and experience chronic pain. Additionally, these individuals with public health cover in Ireland often face substantial delays in accessing specialist care and interventions to manage painful conditions, such as joint replacement, resulting in long-term use of pharmacological treatments. Enhancing access to non-pharmacological interventions may help address reliance on opioids, gabapentinoids and other analgesics, and address medication-related harm, however currently timely access to evidence-based non-pharmacological treatments is limited with geographical variation in service availability.^1717^

Our study appears to be the first to evaluate the unintended consequences of a policy to restrict use of costly lidocaine plasters. We assessed the policy impact from multiple perspectives, including population level analgesic use and from the perspective of individual patients prescribed these products. This study uses dispensing data, which may more closely reflect use than prescribing data, however it is still possible that individuals may not have used medications they had dispensed. The analysis was limited to those on the GMS scheme, which overrepresents older people and more socioeconomically deprived individuals, and we did not include other schemes such as the DPS as pharmacy claims are only captured within the database for households that have prescription spending exceeding the monthly threshold, meaning inconsistent capture for individuals on fewer or less expensive medicines. Individuals may have paid privately for lidocaine plasters after the reimbursement restrictions, which was not captured within the data used in this study. However, given the income threshold for eligibility to the GMS scheme, and low prescription charge (€2.50 per item per month) on the GMS scheme relative to the cost of for private dispensing (in excess of €100), it is unlikely this was a significant pattern among the GMS scheme population. The results may also have been influenced by altered December dispensing behaviour due to the holiday season, however this was mitigated by the PCRS’s policy of limiting each dispensing to one month’s supply and our consideration of a three-month outcome period. We also could not capture over-the-counter medication use, and so some individuals may have purchased non-prescription analgesics, such as ibuprofen, topical diclofenac, paracetamol or co-codamol. Without patient clinical information or patient-reported outcome measures, we could not assess whether the increase in use of other analgesics was clinically appropriate.

### Implications

This study indicates that while the policy change achieved its intended goal of reducing lidocaine plaster utilisation and resultant expenditure, there were significant unintended consequences with regard to increasing utilisation of alternative analgesic agents, particularly opioids and potentially gabapentinoids. For every 1,000 regular lidocaine users before the policy, approximately 133 individuals had opioid treatment intensification, with 87 strong opioid and 26 weak opioid initiations. These consequences have not been previously reported, despite the policy change being implemented approximately eight years ago at the time of publication. Future efforts to reduce low-value care should draw on the growing evidence base for de-implementing by considering multi-level strategies and monitoring negative consequences.^35,36^ Prospective evaluation and monitoring of potential unintended consequences of future pharmaceutical policy changes should be considered, in order to identify and address such impacts in a timely manner. This can be facilitated through the availability and access to data necessary to conduct such evaluations. Determining whether this policy change provided a net benefit requires a comprehensive assessment of these effects, weighing up savings in costs versus relative efficacy and safety of increased use of other analgesics from a relevant decision-maker perspective. Ensuring comprehensive assessment of such policies, incorporating various outcomes, can help balance impacts in terms of effectiveness, safety, and cost-effectiveness, and ultimately maximise benefit to the population.

## Data availability statement

Data were requested from the PCRS for this study through submission of an information request, in line with the PCRS privacy notice (https://www.mymedicalcard.ie/privacy-statement) and procedures, and data were provided to the research team under a data transfer agreement. These procedures were approved by the RCSI University of Medicine and Health Sciences Research Ethics Committee (ref: REC202201015), the Health Service Executive (HSE) Reference Research Ethics Committee B (ref: RRECB1022FM), and the Health Research Consent Declaration Committee (22-008-AF1).

