## Supplementary for "Evaluating the impact of a policy restricting reimbursement of lidocaine plaster prescribing on opioid and other analgesic prescribing"

**Supplementary materials**

Supplementary Table 1. Quantitative results for interrupted time series analysis of lidocaine plasters.

|  | Prevalence of use | | | | Chronic use | | | | Initiations | | | |
| --- | --- | --- | --- | --- | --- | --- | --- | --- | --- | --- | --- | --- |
|  | b | ci95 |  | p | b | ci95 |  | p | b | ci95 |  | p |
| Slope pre-Sept 2017 | 2.20 | 2.14 | 2.27 | <0.001 | 0.81 | 0.78 | 0.83 | <0.001 | 0.56 | 0.51 | 0.61 | <0.001 |
| Level change Sept 2017 | -46.24 | -48.26 | -44.21 | <0.001 | -5.56 | -6.61 | -4.52 | <0.001 | -32.59 | -33.81 | -31.37 | <0.001 |
| Slope change Sept 2017 | -18.30 | -19.04 | -17.56 | <0.001 | -8.06 | -8.56 | -7.56 | <0.001 | -0.31 | -0.37 | -0.25 | <0.001 |
| Level change Dec 2017 | -41.75 | -43.74 | -39.76 | <0.001 | -20.21 | -21.43 | -19.00 | <0.001 | -3.33 | -3.83 | -2.83 | <0.001 |
| Slope change Dec 2017 | 16.38 | 15.64 | 17.12 | <0.001 | 7.46 | 6.96 | 7.95 | <0.001 | -0.29 | -0.33 | -0.24 | <0.001 |
| Intercept | 44.77 | 43.21 | 46.33 | <0.001 | 18.10 | 17.61 | 18.60 | <0.001 | 15.56 | 14.87 | 16.25 | <0.001 |

Supplementary Table 2. Quantitative results for interrupted time series analysis of other analgesic drugs

|  | **Prevalence** | | | | **Chronic use** | | | | **Initiations** | | | |
| --- | --- | --- | --- | --- | --- | --- | --- | --- | --- | --- | --- | --- |
| **Topical NSAIDs** | b | ci95 |  | p | b | ci95 |  | p | b | ci95 |  | p |
| Slope pre-Sept 2017 | 1.67 | 1.44 | 1.90 | 0.000 | 0.58 | 0.51 | 0.65 | 0.000 | 0.83 | 0.67 | 1.00 | 0.000 |
| Level change Sept 2017 | 10.65 | 2.12 | 19.19 | 0.014 | 0.42 | -1.27 | 2.12 | 0.624 | 2.01 | -3.65 | 7.68 | 0.486 |
| Slope change Sept 2017 | 0.96 | -3.03 | 4.94 | 0.638 | 0.47 | 0.01 | 0.93 | 0.046 | -1.91 | -4.61 | 0.78 | 0.164 |
| Level change Dec 2017 | -5.42 | -17.61 | 6.76 | 0.383 | 1.98 | -1.13 | 5.08 | 0.213 | -0.97 | -9.87 | 7.93 | 0.831 |
| Slope change Dec 2017 | -0.94 | -4.96 | 3.08 | 0.646 | -0.26 | -0.77 | 0.25 | 0.314 | 1.22 | -1.51 | 3.95 | 0.381 |
| Intercept | 258.41 | 252.98 | 263.85 | 0.000 | 69.53 | 67.87 | 71.19 | 0.000 | 82.67 | 79.35 | 86.00 | 0.000 |
| **Topical capsaicin** | b | ci95 |  | p | b | ci95 |  | p | b | ci95 |  | p |
| Slope pre-Sept 2017 | 0.02 | 0.02 | 0.03 | 0.000 | 0.00 | 0.00 | 0.01 | 0.001 | 0.02 | 0.01 | 0.02 | 0.000 |
| Level change Sept 2017 | 1.54 | 1.21 | 1.88 | 0.000 | -0.02 | -0.15 | 0.12 | 0.827 | 1.45 | 1.21 | 1.68 | 0.000 |
| Slope change Sept 2017 | 0.54 | 0.34 | 0.74 | 0.000 | 0.03 | -0.05 | 0.11 | 0.510 | 0.11 | -0.03 | 0.24 | 0.116 |
| Level change Dec 2017 | 0.89 | -0.02 | 1.79 | 0.054 | 0.68 | 0.34 | 1.03 | 0.000 | -0.04 | -0.46 | 0.38 | 0.839 |
| Slope change Dec 2017 | -0.74 | -0.96 | -0.52 | 0.000 | -0.06 | -0.15 | 0.02 | 0.150 | -0.21 | -0.35 | -0.08 | 0.002 |
| Intercept | 4.87 | 4.72 | 5.01 | 0.000 | 1.24 | 1.20 | 1.28 | 0.000 | 1.80 | 1.71 | 1.90 | 0.000 |
| **Opioids** | b | ci95 |  | p | b | ci95 |  | p | b | ci95 |  | p |
| Slope pre-Sept 2017 | 2.00 | 1.68 | 2.33 | 0.000 | 1.37 | 1.22 | 1.51 | 0.000 | 0.79 | 0.59 | 1.00 | 0.000 |
| Level change Sept 2017 | -8.18 | -19.58 | 3.22 | 0.160 | -7.64 | -11.59 | -3.69 | 0.000 | -7.40 | -13.95 | -0.86 | 0.027 |
| Slope change Sept 2017 | 4.58 | -0.80 | 9.96 | 0.095 | 1.00 | -0.58 | 2.58 | 0.216 | 1.62 | -1.40 | 4.65 | 0.293 |
| Level change Dec 2017 | -1.93 | -18.21 | 14.35 | 0.816 | 6.76 | 0.81 | 12.71 | 0.026 | -4.40 | -12.91 | 4.11 | 0.311 |
| Slope change Dec 2017 | -5.02 | -10.43 | 0.40 | 0.069 | -1.23 | -2.84 | 0.39 | 0.136 | -2.37 | -5.41 | 0.67 | 0.126 |
| Intercept | 575.27 | 567.26 | 583.28 | 0.000 | 265.98 | 262.65 | 269.31 | 0.000 | 113.33 | 109.27 | 117.39 | 0.000 |
| **Systemic NSAIDs** | b | ci95 |  | p | b | ci95 |  | p | b | ci95 |  | p |
| Slope pre-Sept 2017 | -0.31 | -0.77 | 0.16 | 0.199 | -0.04 | -0.13 | 0.05 | 0.376 | 1.50 | 1.10 | 1.90 | 0.000 |
| Level change Sept 2017 | -9.79 | -29.67 | 10.08 | 0.334 | -3.51 | -5.72 | -1.30 | 0.002 | -13.69 | -26.33 | -1.05 | 0.034 |
| Slope change Sept 2017 | -0.19 | -11.05 | 10.66 | 0.972 | -0.74 | -1.49 | 0.02 | 0.057 | -2.65 | -8.69 | 3.40 | 0.391 |
| Level change Dec 2017 | 12.22 | -17.25 | 41.69 | 0.416 | 5.70 | 2.82 | 8.59 | 0.000 | 4.71 | -12.19 | 21.62 | 0.585 |
| Slope change Dec 2017 | -0.48 | -11.36 | 10.41 | 0.932 | 0.83 | 0.05 | 1.60 | 0.036 | 0.76 | -5.30 | 6.82 | 0.807 |
| Intercept | 580.06 | 568.16 | 591.95 | 0.000 | 117.01 | 114.86 | 119.17 | 0.000 | 175.97 | 167.74 | 184.19 | 0.000 |
| **Gabapentinoids** | b | ci95 |  | p | b | ci95 |  | p | b | ci95 |  | p |
| Slope pre-Sept 2017 | 2.01 | 1.90 | 2.12 | 0.000 | 1.35 | 1.24 | 1.46 | 0.000 | 0.12 | 0.08 | 0.15 | 0.000 |
| Level change Sept 2017 | -8.76 | -12.45 | -5.07 | 0.000 | -6.67 | -9.59 | -3.75 | 0.000 | -1.27 | -2.28 | -0.25 | 0.015 |
| Slope change Sept 2017 | -0.86 | -2.32 | 0.61 | 0.251 | -0.21 | -1.32 | 0.90 | 0.710 | -0.33 | -0.73 | 0.08 | 0.112 |
| Level change Dec 2017 | 1.30 | -3.44 | 6.03 | 0.591 | 3.97 | -0.11 | 8.06 | 0.057 | -0.72 | -2.25 | 0.82 | 0.360 |
| Slope change Dec 2017 | -0.92 | -2.39 | 0.56 | 0.225 | -0.88 | -2.01 | 0.25 | 0.127 | 0.15 | -0.27 | 0.56 | 0.488 |
| Intercept | 220.00 | 217.54 | 222.47 | 0.000 | 132.23 | 129.91 | 134.54 | 0.000 | 24.26 | 23.50 | 25.02 | 0.000 |
| **Paracetamol** | b | ci95 |  | p | b | ci95 |  | p | b | ci95 |  | p |
| Slope pre-Sept 2017 | 3.41 | 2.78 | 4.05 | 0.000 | 2.00 | 1.73 | 2.26 | 0.000 | 1.34 | 0.94 | 1.75 | 0.000 |
| Level change Sept 2017 | -3.17 | -24.13 | 17.78 | 0.767 | -9.84 | -16.26 | -3.42 | 0.003 | -5.03 | -16.49 | 6.42 | 0.389 |
| Slope change Sept 2017 | 6.49 | -2.14 | 15.13 | 0.141 | 0.90 | -0.93 | 2.74 | 0.336 | -0.25 | -4.79 | 4.29 | 0.914 |
| Level change Dec 2017 | 9.76 | -23.47 | 42.99 | 0.565 | 15.49 | 6.57 | 24.40 | 0.001 | 0.14 | -17.39 | 17.68 | 0.987 |
| Slope change Dec 2017 | -6.60 | -15.38 | 2.19 | 0.141 | -0.66 | -2.59 | 1.26 | 0.498 | -1.05 | -5.66 | 3.56 | 0.656 |
| Intercept | 886.80 | 871.31 | 902.29 | 0.000 | 349.00 | 342.95 | 355.06 | 0.000 | 172.73 | 164.61 | 180.85 | 0.000 |

Supplementary Table 3. Summary of absolute risk increases and number needed to treat for each binary outcome.

|  | Control event rate* | RR (95%CI) | | | ARI (95%CI) | | | NNT (95%CI) | | |
| --- | --- | --- | --- | --- | --- | --- | --- | --- | --- | --- |
| **Treatment intensification** |  |  | | |  | | |  | | |
| Opioids | 28.3 | 1.47 | 1.38 | 1.57 | 0.133 | 0.108 | 0.161 | 7.5 | 6.2 | 9.3 |
| Gabapentinoids | 12.1 | 1.26 | 1.16 | 1.38 | 0.031 | 0.019 | 0.046 | 31.8 | 21.7 | 51.7 |
| Systemic NSAIDs | 20.1 | 1.12 | 1.04 | 1.21 | 0.024 | 0.008 | 0.042 | 41.5 | 23.7 | 124.4 |
| Paracetamol | 34.7 | 1.35 | 1.27 | 1.44 | 0.121 | 0.094 | 0.153 | 8.2 | 6.5 | 10.7 |
| **Initiation** |  |  |  |  |  |  |  |  |  |  |
| Strong opioids | 8.1 | 2.07 | 1.85 | 2.3 | 0.087 | 0.069 | 0.105 | 11.5 | 9.5 | 14.5 |
| Weak opioids | 6.9 | 1.38 | 1.21 | 1.59 | 0.026 | 0.014 | 0.041 | 38.1 | 24.6 | 69.0 |
| Gabapentinoids | 3.5 | 1.34 | 1.11 | 1.61 | 0.012 | 0.004 | 0.021 | 84.0 | 46.8 | 259.7 |
| Systemic NSAIDs | 13.6 | 1.13 | 1.02 | 1.26 | 0.018 | 0.003 | 0.035 | 56.6 | 28.3 | 367.6 |
| Paracetamol | 25 | 1.66 | 1.49 | 1.86 | 0.165 | 0.123 | 0.215 | 6.1 | 4.7 | 8.2 |

*Indicates the percentage of control group patients (i.e. users of other analgesics without lidocaine plasters pre-policy implementation) experiencing each outcome. ARI = absolute risk increase; NNT = number needed to treat, i.e. the number of individuals to be exposed to the policy of lidocaine plaster reimbursement restriction for one additional case of treatment intensification or initiation to occur; NSAID = non-steroidal anti-inflammatory drug; RR = risk ratio

Supplementary Table 4. Adjusted regression analyses stratified by age group.

|  | Aged >=65 years | Aged <65 years |
| --- | --- | --- |
| **Treatment intensification (Adjusted RR, 95%CI)** |  |  |
| Opioids | 1.46 (1.35-1.57) | 1.49 (1.32-1.68) |
| Gabapentinoids | 1.29 (1.16-1.44) | 1.18 (1.01-1.38) |
| Systemic NSAIDs | 1.10 (0.99-1.21) | 1.20 (1.05-1.37) |
| Paracetamol | 1.36 (1.27-1.47) | 1.32 (1.16-1.49) |
| **Initiation (Adjusted RR, 95%CI)** |  |  |
| Strong opioids | 2.07 (1.83-2.34) | 2.06 (1.64-2.60) |
| Weak opioids | 1.40 (1.19-1.64) | 1.34 (1.02-1.77) |
| Gabapentinoids | 1.38 (1.22-1.71) | 1.17 (0.79-1.72) |
| Systemic NSAIDs | 1.09 (0.96-1.23) | 1.23 (1.03-1.47) |
| Paracetamol | 1.78 (1.56-2.02) | 1.40 (1.13-1.73) |
| **Rate of standard doses (Adjusted IRR, 95%CI)** |  |  |
| Opioids | 1.09 (1.05-1.14) | 1.22 (1.14-1.30) |
| Gabapentinoids | 0.97 (0.94-1.01) | 1.01 (0.97-1.06) |
| Systemic NSAIDs | 1.05 (1.01-1.10) | 1.01 (0.96-1.06) |
| Paracetamol | 1.08 (1.06-1.09) | 1.05 (1.02-1.09) |

CI = confidence interval; IRR = incidence rate ratio; NSAID = non-steroidal anti-inflammatory drug; RR = risk ratio

Supplementary Table 5. Sensitivity analysis adjusting for difference between treatment group in November 2016

|  | **Adjusted RR** | **95% CI** | |
| --- | --- | --- | --- |
| **Treatment intensification** |  |  |  |
| Opioids | 1.18 | 1.09 | 1.28 |
| Gabapentinoids | 0.99 | 0.89 | 1.10 |
| Systemic NSAIDs | 1.23 | 1.11 | 1.36 |
| Paracetamol | 1.14 | 1.06 | 1.24 |
| **Initiation** |  |  |  |
| Strong opioids | 1.22 | 1.05 | 1.42 |
| Weak opioids | 1.22 | 1.00 | 1.49 |
| Gabapentinoids | 0.99 | 0.77 | 1.27 |
| Systemic NSAIDs | 1.25 | 1.09 | 1.44 |
| Paracetamol | 1.04 | 0.90 | 1.22 |
| **Rate of standard doses*** |  |  |  |
| Opioids | 0.99 | 0.94 | 1.04 |
| Gabapentinoids | 1.03 | 0.99 | 1.07 |
| Systemic NSAIDs | 1.06 | 1.01 | 1.11 |
| Paracetamol | 1.00 | 0.98 | 1.02 |

CI = confidence interval; NSAID = non-steroidal anti-inflammatory drug; RR = risk ratio

* Adjusted incidence rate ratio

**Supplementary Table 6. Sensitivity analysis including alll regression results for individuals dispensed lidocaine in November 2017 versus individuals dispensed other analgesics**

|  | **Adjusted RR** | **95% CI** | |
| --- | --- | --- | --- |
| **Treatment intensification** |  |  |  |
| Opioids | 1.49 | 1.44 | 1.56 |
| Gabapentinoids | 1.33 | 1.26 | 1.42 |
| Systemic NSAIDs | 1.09 | 1.04 | 1.15 |
| Paracetamol | 1.37 | 1.31 | 1.43 |
| **Initiation** |  |  |  |
| Strong opioids | 1.88 | 1.74 | 2.02 |
| Weak opioids | 1.33 | 1.22 | 1.45 |
| Gabapentinoids | 1.50 | 1.34 | 1.68 |
| Systemic NSAIDs | 1.08 | 1.01 | 1.15 |
| Paracetamol | 1.58 | 1.47 | 1.69 |
| **Rate of standard doses*** |  |  |  |
| Opioids | 1.12 | 1.09 | 1.15 |
| Gabapentinoids | 0.99 | 0.97 | 1.01 |
| Systemic NSAIDs | 1.06 | 1.04 | 1.09 |
| Paracetamol | 1.09 | 1.07 | 1.10 |

CI = confidence interval; NSAID = non-steroidal anti-inflammatory drug; RR = risk ratio

* Adjusted incidence rate ratio

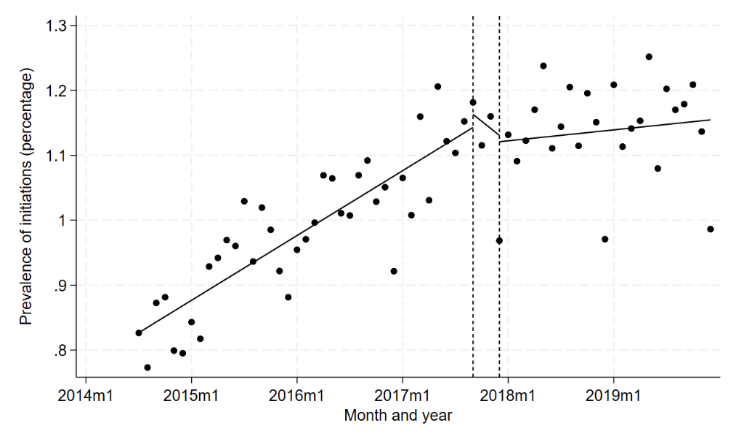

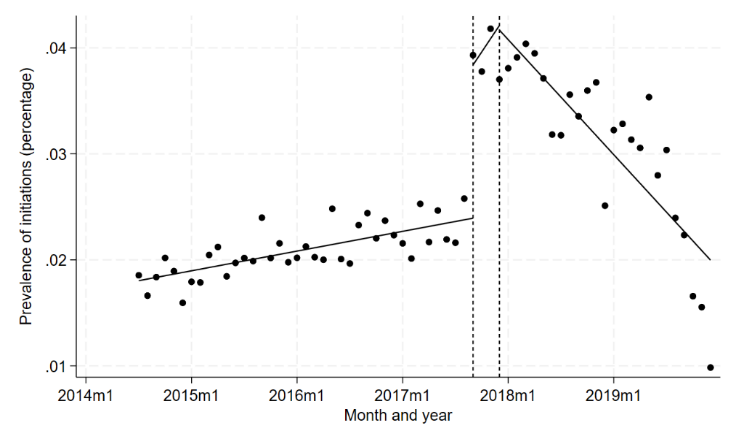

D

B

A

C

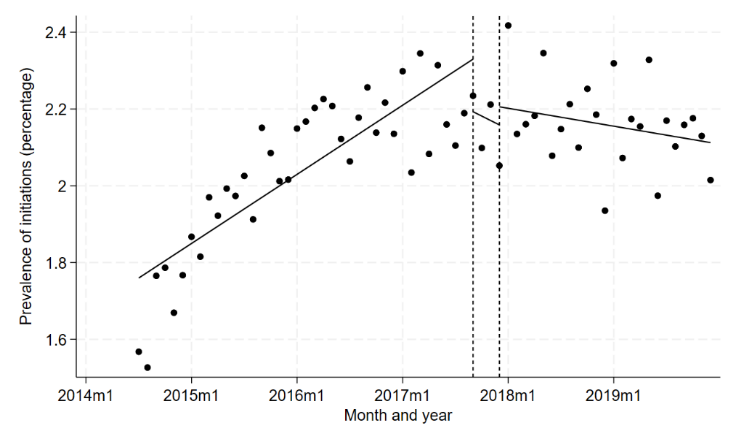

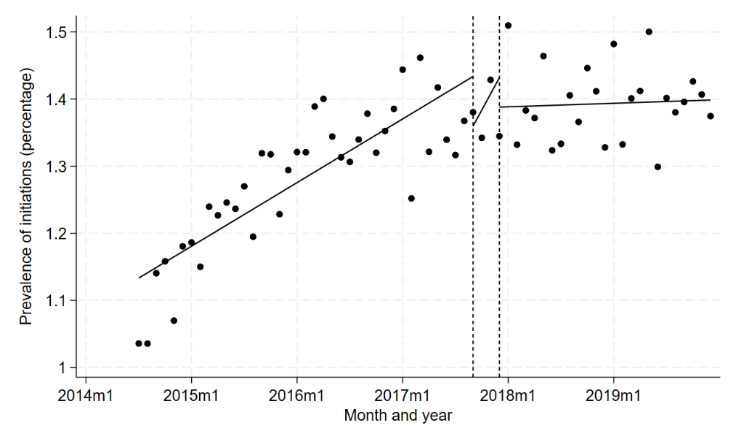

E

FF

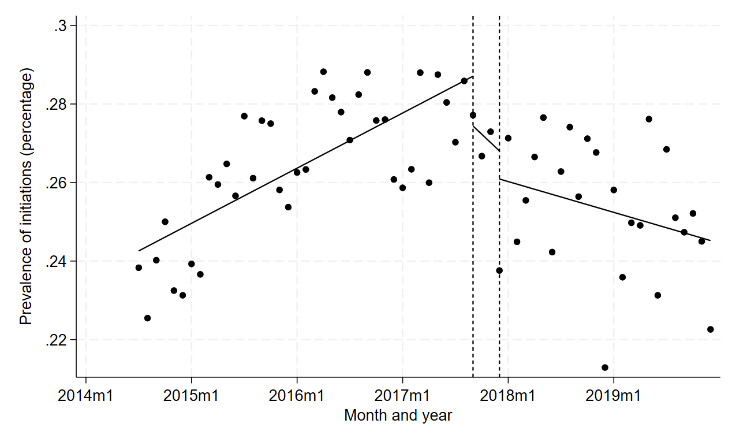

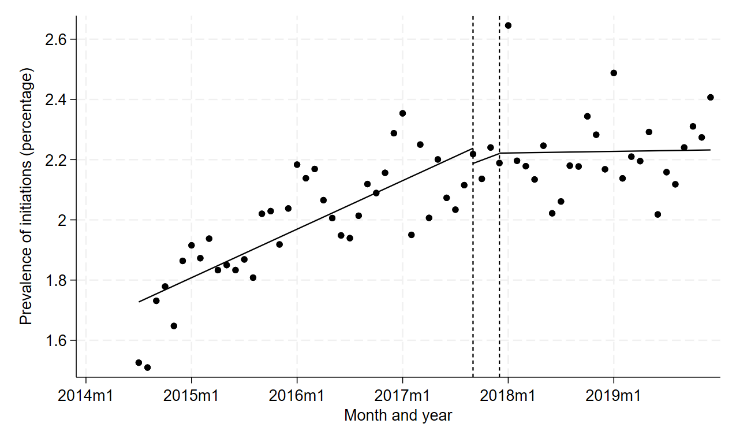

Supplementary Figure 1. Interrupted time series analysis for initiations of (a) topical NSAIDs, (b) topical capsaicin, (c) systemic NSAIDs, (d) opioids, (e) gabapentinoids, and (f) paracetamol.

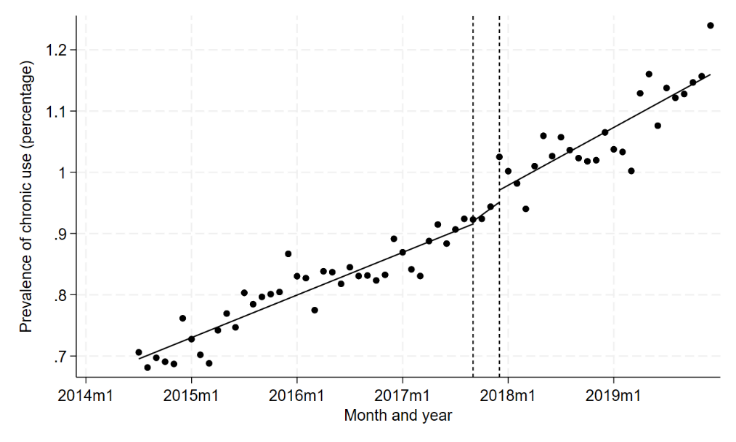

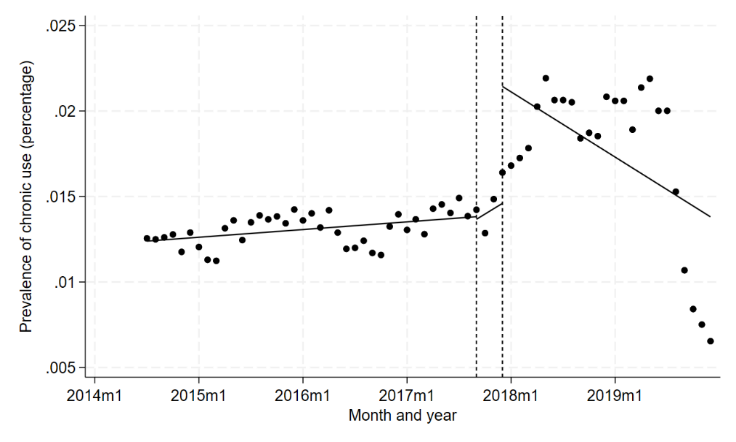

B

D

A

C

F

E

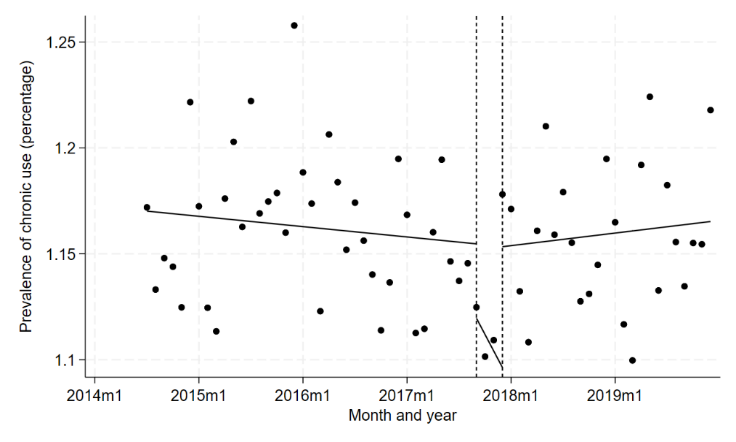

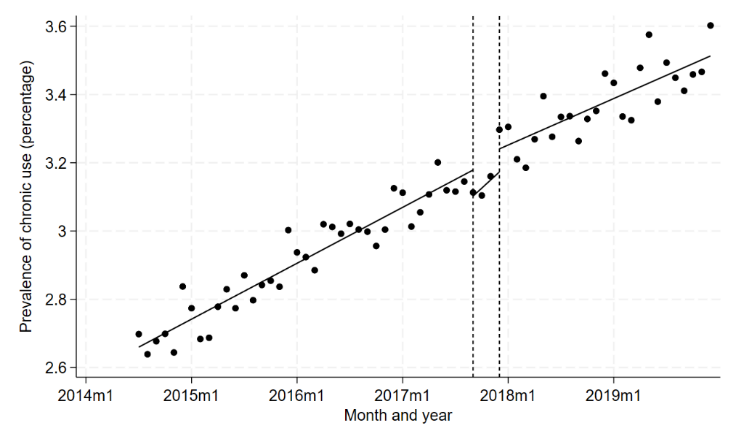

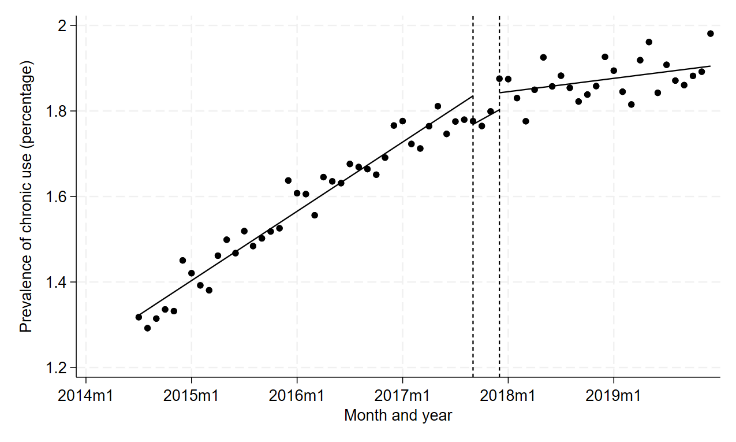

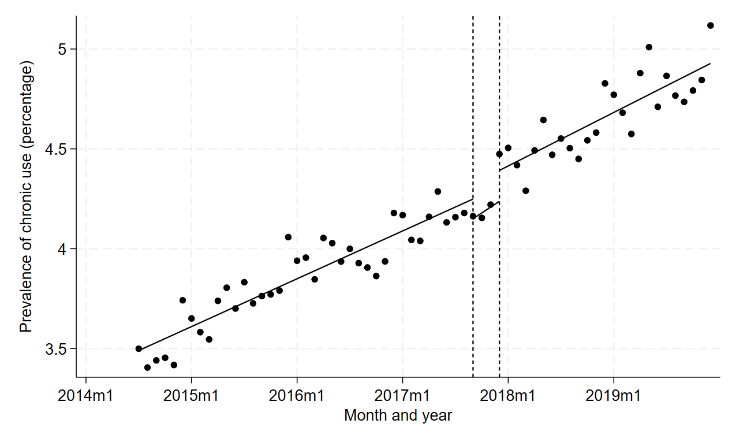

Supplementary Figure 2. Interrupted time series analysis for chronic use of (a) topical NSAIDs, (b) topical capsaicin, (c) systemic NSAIDs, (d) opioids, (e) gabapentinoids, and (f) paracetamol.
